# Eating disorder symptoms and their associations with anthropometric and psychiatric polygenic scores

**DOI:** 10.1101/2021.06.02.21258214

**Authors:** Mohamed Abdulkadir, Christopher Hübel, Moritz Herle, Ruth J.F. Loos, Gerome Breen, Cynthia M. Bulik, Nadia Micali

## Abstract

**Background:** Eating disorder (ED) symptoms are prevalent in the general population, but their shared genetic underpinnings with psychiatric, metabolic, and anthropometric traits are not known. Here, we examined if polygenic scores (PGS) of traits associated with anorexia nervosa are also associated with adolescent ED symptoms in the Avon Longitudinal Study of Parents and Children (ALSPAC).

**Methods:** We calculated PGS for 25 traits (16 psychiatric, 4 metabolic, and 5 anthropometric) and investigated their associations with eight ED symptoms, including behaviors such as fasting for weight loss and cognitions such as body dissatisfaction (n range=2,991-6,013).

**Results:** Higher attention deficit hyperactivity disorder PGS and lower educational attainment PGS were associated with fasting for weight loss. Higher insomnia PGS was associated with increased body dissatisfaction. We found no evidence of an association between metabolic trait PGS and any ED symptom. Regarding anthropometrics, fat-free mass, fat mass, and body fat percentage PGSs, were positively associated with binge eating, excessive exercise, fasting for weight loss, body dissatisfaction, and weight and shape concern.

**Conclusions:** ED symptoms are genetically associated with psychiatric and anthropometric, but not with metabolic traits. Our findings provide insights for future genetic research investigating why some individuals with ED symptoms progress to develop threshold EDs while others do not.

## Introduction

Eating disorder (ED) symptoms are prevalent (18%-22%) in the general population and include behavioral symptoms (e.g., binge eating, purging, restricting) and cognitive symptoms (e.g., body dissatisfaction, drive for thinness) (Hayes et al., 2018; Micali et al., 2015, 2017). Epidemiological studies suggest that ED symptoms are phenotypically associated with psychiatric traits (e.g., mood disorders, psychosocial impairment, and suicidal behavior) and anthropometric traits (Chamay-Weber, Narring, & Michaud, 2005; Crow, Eisenberg, Story, & Neumark-Sztainer, 2008; Micali et al., 2017; Reed, Micali, Bulik, Smith, & Wade, 2017). Especially for anthropometric traits, recent evidence suggests a causal bidirectional relationship between ED symptoms and body mass index (BMI): higher BMI has an effect on increased ED symptoms and vice versa (Reed et al., 2017). Therefore, given the continuing rising prevalence of obesity worldwide (Jaacks et al., 2019), the prevalence of eating disorder symptoms may rise in tandem. From a public health perspective, it is therefore important to understand the etiology of ED symptoms (Dinkler et al., 2019).

ED symptoms have been shown to be moderately heritable (*<u>h^2^</u>* = 0.65, 95% confidence interval = 0.62–0.69; Dinkler et al., 2019). However, the genetic architecture of ED symptoms has received little attention. To date, only two genome-wide association studies (GWASs; a genetic study in which genetic variants, across the entire genome, are tested for their association with a trait of interest) of ED symptoms (e.g., drive for thinness, body dissatisfaction, and bulimia) (Boraska et al., 2012; Wade et al., 2013). Although no genome-wide significant loci were detected, the authors did report that the top single nucleotide polymorphisms (SNPs; single base-pair difference in the DNA sequence of among individuals in a population) had previously been associated with other psychiatric disorders, namely major depressive disorder and schizophrenia (Boraska et al., 2012). The lack of genome-wide significant signal was likely due to the typically small-to-modest effect sizes of the common SNPs that influence complex traits that require large sample sizes in the thousands for detection (Boraska et al., 2012).

Polygenic score (PGS) analysis is a powerful method that leverages the small-to-modest effect sizes of common SNPs into a single continuous score that exceeds the explanatory power of single SNPs (Purcell et al., 2009). PGSs can be used to examine whether two traits are genetically related by associating the PGSs calculated for one trait with another trait. Our group (Abdulkadir et al., 2020) and others (Nagata et al., 2019; Robinson et al., 2020; Solmi, Mascarell, Zammit, Kirkbride, & Lewis, 2019) have successfully applied this method and reported that a BMI PGS and a schizophrenia PGS are associated with eating disorder symptoms (e.g., binge eating, fasting for weight loss, and body dissatisfaction) in the general population. However, a broader evaluation of PGSs indexing psychiatric and anthropometric/metabolic traits is warranted, as a GWAS on anorexia nervosa (AN) reported several significant genetic correlations with psychiatric, metabolic, and anthropometric traits (Duncan et al., 2017; Hübel et al., 2019; Watson et al., 2019).

The aim of this study was to investigate whether PGSs of psychiatric, metabolic, and anthropometric traits that are genetically associated with AN are also associated with behavioral and cognitive ED symptoms in the Avon Longitudinal Study of Parents and Children (ALSPAC). We hypothesized that like AN (Watson et al., 2019), behavioral and cognitive ED symptoms would be associated with anthropometric, metabolic, and psychiatric PGSs.

## Methods

### Participants

The ALSPAC study is an ongoing population-based birth cohort study of mothers and their children (that were born between 1 April 1991 and 31 December 1992) residing in the southwest of England (UK) (Boyd et al., 2013; Fraser et al., 2013; Golding, 2004; Golding, Pembrey, Jones, & The Alspac Study Team, 2001; Northstone et al., 2019). From the 14,541 pregnancies, 13,988 were alive at 1 year. At age 7 years, this sample was bolstered with an additional 913 children. The total sample size for analyses using any data collected after the age of seven is therefore 15,454 pregnancies; of these 14,901 were alive at 1 year of age. Participants are assessed at regular intervals using clinical interviews, self-report questionnaires, medical records, and physical examinations. Please note that the study website contains details of all the data that are available through a fully searchable data dictionary and variable search tool and reference the following webpage: http://www.bristol.ac.uk/alspac/researchers/our-data/. To avoid potential confounding due to relatedness, one sibling per set of multiple births was randomly selected to guarantee independence of participants (*N* = 75). Furthermore, individuals who were closely related to each other (defined as a φ > 0.2, as calculated in PLINK v1.90b), were removed.

### Ethics statement

The authors assert that all procedures contributing to this work comply with the ethical standards of the relevant national and institutional committees on human experimentation and with the Helsinki Declaration of 1975, as revised in 2008. Ethical approval for the study was obtained from the ALSPAC Ethics and Law Committee and the Local Research Ethics Committees (Bristol and Weston Health Authority: E1808 Children of the Nineties: ALSPAC, 28th November 1989 (for details see: www.bristol.ac.uk/alspac/researchers/research-ethics/).

Consent for biological samples was obtained in accordance with the Human Tissue Act (2004). Informed consent for the use of data collected via questionnaires and clinics was obtained from participants following the recommendations of the ALSPAC Ethics and Law Committee at the time. The main caregiver initially provided consent for child participation and from the age 16 years the offspring themselves have provided informed written consent.

### ED symptoms: behaviors

Excessive exercise, fasting for weight loss, binge eating, and purging were assessed at age 14, 16, and 18 years using adapted questionnaire items from the Youth Risk Behavior Surveillance System (Kann et al., 1996), which has previously been validated in a population-based study (Field, Taylor, Celio, & Colditz, 2004).

#### Excessive exercise

Individuals who endorsed exercising for weight loss purposes and felt guilty about missing exercise and found it hard to meet other obligations, such as schoolwork, because of their exercise regime at any of the ages assessed (14, 16, or 18 years) were coded as engaging in excessive exercise.

#### Fasting for weight loss

Participants reported the frequency of fasting (i.e., not eating for an entire day) to lose or avoid gaining weight during the past year. Similar to the excessive exercise variable, individuals that indicated any fasting for weight loss at any age (14, 16, or 18 years) were coded as engaging in fasting for weight loss.

#### Binge eating

Participants reported the frequency of binge eating during the past year—defined as eating an unusually large amount of food, combined with a feeling of being out of control. Those who reported having engaged in binge eating at either age 14, 16, or 18 years were coded as engaging in binge eating.

#### Purging

Participants reported the frequency with which they used laxatives or self-induced vomiting, or other medicines to lose or avoid gaining weight during the past year; individuals that responded affirmatively to all of these questions at either the age 14, 16, or 18 years were coded as engaging in purging.

### ED symptoms: cognitions

Fear of weight gain and weight and shape concern were also assessed with adapted questionnaire items from the Youth Risk Behavior Surveillance System (Kann et al., 1996).

#### Fear of weight gain

At age 13, 14 and 16 years, parents reported if their child was afraid of gaining weight or getting fat (0 = “No”, 1 = “A little”, 2 = “A lot”, and 3 = “It really terrified him/her”), if they avoided fattening foods (0 = “No”, 1 = “A little”, 2 = “A lot”) or whether their child felt it would be difficult or impossible if they were asked to put on 2 kilos for the sake of their health (0 = “Easy”, 1 = “Difficult”, 2 = “Impossible”). The maximum value of either of the three ages (13, 14, and 16 years) was used for the analyses; a higher score indicates greater fear of weight gain.

#### Thin ideal internalization

At age 14 years, thin ideal internalization was assessed using the Ideal-Body Stereotype Scale-Revised (Stice, 1998) with gender-specific items used to assess different aspects of appearance-ideal internalization for girls and boys separately. Adolescent girls and boys reported how strongly they agreed a series of questions; e.g., thin women being not attractive and thin men being more good-looking. The responses were rated on a five-point Likert scale from “strongly agree” to “strongly disagree” and the items from this scale were summed to obtain a total score; a higher score corresponded with increased internalization of the thin ideal.

#### Weight and shape concern

At age 14 years, weight and shape concerns were assessed by using questions from the McKnight Risk Factor Survey (Shisslak et al., 1999). Participants were asked how they felt about (i) the way their body looked, (ii) how much their weight had made a difference to how they feel about themselves, and (iii) how much they had worried about gaining a little weight (e.g., 1 kg). Responses were captured on a four response Likert scale ranging from “very happy” or “not at all” to “very unhappy” or “a lot”. These three questions were summed to generate an overall score.

#### Body dissatisfaction

Body dissatisfaction was assessed at age 14 years using the Body Dissatisfaction Scale (Calzo, Austin, & Micali, 2018; Micali et al., 2015; Stice, 2001). Participants were asked gender-specific questions rating their satisfaction with nine body parts with responses on a six-point Likert scale ranging from ‘extremely satisfied’ to ‘extremely dissatisfied’. We constructed a continuous score from this scale with a higher score corresponding to higher body dissatisfaction.

### Body mass index

BMI (weight/height^2^) was calculated using objectively measured weight and height obtained during a face-to-face assessment at age 11 years (Boyd et al., 2013, 2019; Fraser et al., 2013; Northstone et al., 2019). Height was measured to the nearest millimeter using a Harpenden Stadiometer (Holtain Ltd., Crymych, UK) and weight was measured using the Tanita Body Fat analyzer (Tanita TBF UK Ltd., Middlesex, UK) to the nearest 50 g. Age- and sex-standardized BMI z-scores (zBMI) were calculated according to UK reference data, indicating the degree to which a child is heavier (>0) or lighter than expected according to his/her age and sex (Cole, Bellizzi, Flegal, & Dietz, 2000).

### Selection of PGS

PGSs index an individual’s genetic liability to a trait. They are calculated by summing the genetic variants an individual carries and weighting these genetic variants by the corresponding effect sizes that have been detected in previous independent GWASs. Using GWAS summary statistics from independent samples, we calculated PGSs for 25 traits that we grouped into three broad categories; (i) PGS from psychiatric/neurological/behavioral traits and one PGS indexing educational attainment (*N*_PGS_ = 16); (ii) metabolic traits (*N*_PGS_ = 4); and (iii) anthropometric traits (*N*_PGS_ = 5). In the first category, the selection of the psychiatric PGSs was partially based on the previously reported significant genetic correlations of AN with these traits (Watson et al., 2019): schizophrenia (Ripke et al., 2014), post-traumatic stress disorder (Duncan et al., 2018), obsessive-compulsive disorder (OCD) (Arnold et al., 2018), neuroticism (Hübel et al., 2019), major depressive disorder (Wray et al., 2018), and lifetime probable anxiety disorder (Purves et al., 2020). To test the hypothesis whether ED symptoms are related to a broader array of psychiatric traits we also included PGSs for lifetime cannabis use (Pasman et al., 2018), insomnia (Jansen et al., 2019), borderline personality disorder (Witt et al., 2017), bipolar disorder (Stahl et al., 2019), autism spectrum disorder (Grove et al., 2019), attention deficit hyperactivity disorder (ADHD) (Demontis et al., 2019), AN (Watson et al., 2019), and alcohol dependence (Walters et al., 2018). We also calculated PGS for physical activity (Hübel et al., 2019), migraine (Gormley et al., 2016), and educational attainment (Lee et al., 2018) as previous research suggests an association with AN (Watson et al., 2019). In the second category, the selection of metabolic traits was also partially based on the previously identified genetic correlations with AN (Watson et al., 2019): type 2 diabetes (Scott et al., 2017), insulin resistance (Scott et al., 2017), fasting insulin (Dupuis et al., 2010), and high-density lipoprotein (HDL) cholesterol (Teslovich et al., 2010). Lastly, the third category included the anthropometric traits of height (Yengo et al., 2018), fat-free mass, fat mass, and body fat percentage (Hübel et al., 2019); all of these traits were previously associated with either AN or ED symptoms (Hübel et al., 2019; Watson et al., 2019). For a full list of PGS included in this study, see **Supplementary Table S1.**

### Polygenic risk scoring

We used PRSice version 2.2.3 for calculating the PGSs (Choi & O’Reilly, 2019). We clumped the SNPs that were present both in the summary statistics of the trait and in the genotype data of the ALSPAC (i.e., overlapping SNPs) to obtain genetically independent SNPs. We retained the SNP with the smallest *p* value in each 250 kilobase window of all those in linkage disequilibrium (*r^2^* > 0.1). We calculated PGS at their optimal *p* value threshold in each individual by scoring the number of effect alleles (weighted by the allele effect size) across the set of remaining SNPs. We calculated the PGS using the high-resolution scoring (i.e., incrementally across a large number of *p* value thresholds) method to identify the *p* value threshold at which the PGS is optimally associated with the outcome and explains the most variance (i.e., resulting in the highest adjusted *R^2^*for continuous outcomes and Nagelkerke’s *R^2^*on the liability scale for binary outcomes). We evaluated the associations between PGS and ED symptoms using generalized linear models, adjusted for sex and the first six principal components. To adjust for overfitting, we permuted case-control status at every *p* value threshold 10,000 times and calculated empirical *p* values. For the associations between the PGS and the binary ED symptoms (i.e., binge eating, excessive exercise, fasting, fear of weight gain, and purging) we report odds ratios (OR) and for the associations between the polygenic scores and the and the continuous ED symptoms (body dissatisfaction, thin ideal internalization, and weight and shape concerns) we report beta coefficients. To correct for multiple testing (i.e., 25 PGS regression models), we calculated *Q* values using the false discovery rate approach (Benjamini & Hochberg, 1995).

### Exploratory causal mediation analyses

In previous work, we demonstrated that BMI is associated with ED symptoms in ALSPAC (Abdulkadir et al., 2020) and given reports that psychiatric disorders and educational attainment are associated with BMI (Benson, von Hippel, & Lynch, 2018; Hübel et al., 2019; McCrea, Berger, & King, 2012), it is therefore possible that association between psychiatric and the education-related PGS with an ED symptoms could be mediated through measured BMI. As a first step, in the case of a significant association in the main PGS analyses (described above), the psychiatric and the education-related PGS were tested for an association with BMI at age 11 years (before endorsement of the ED symptoms). Only the PGS that showed both a significant association with an ED symptom and BMI at age 11 years were included in the mediation analyses to estimate the natural direct and indirect effect using concepts proposed in modern causal inference; the natural direct and indirect effects (VanderWeele, 2015). The natural direct effect (also known as the “average direct effect”) measures the mean or expected risk difference (in the case of a binary outcome measure) had the PGS been hypothetically set to change by 1 SD from 0 to 1, while at the same time childhood BMI had been set to take its natural value (i.e., the value that would be experienced had the PGS been set at the reference value of 0). The natural indirect effect (also known as the average causal mediation effect) measures the expected mean difference or expected risk difference (in the case of a binary outcome measure) had the PGS been hypothetically set to take the value 1, while at the same time childhood BMI had been set to take its potential values had PGS been set to 0 or 1. When summed, these direct and indirect effects give the total causal effects and therefore are useful measures of the contribution made by a particular pathway to a causal relationship. The mediation analyses were carried out using the ‘mediate’ package in R (Tingley, Yamamoto, Hirose, Keele, & Imai, 2014). Analyses were controlled for biological sex, and the first six ancestry-informative principal components. Furthermore, we assumed that there were no additional unaccounted confounders nor any intermediate confounders (VanderWeele, 2015).

### Results

#### Sample description

Following quality control of the genetic data, between 2,991 and 5,225 individuals remained eligible for analyses with both genotype and phenotype data (Table 1-2). We observed weak to moderate correlations (*r* = 0.13–0.72; Figure 1) among the ED symptom variables with a notable exception for thin ideal internalization that showed negligible correlations with any of the ED symptoms (*r* = -0.01–0.06). Thin idealization also differed from the other ED symptoms as it was the only ED symptom that showed consistent negative correlation with BMI at age 11 years.

**Figure 1:**
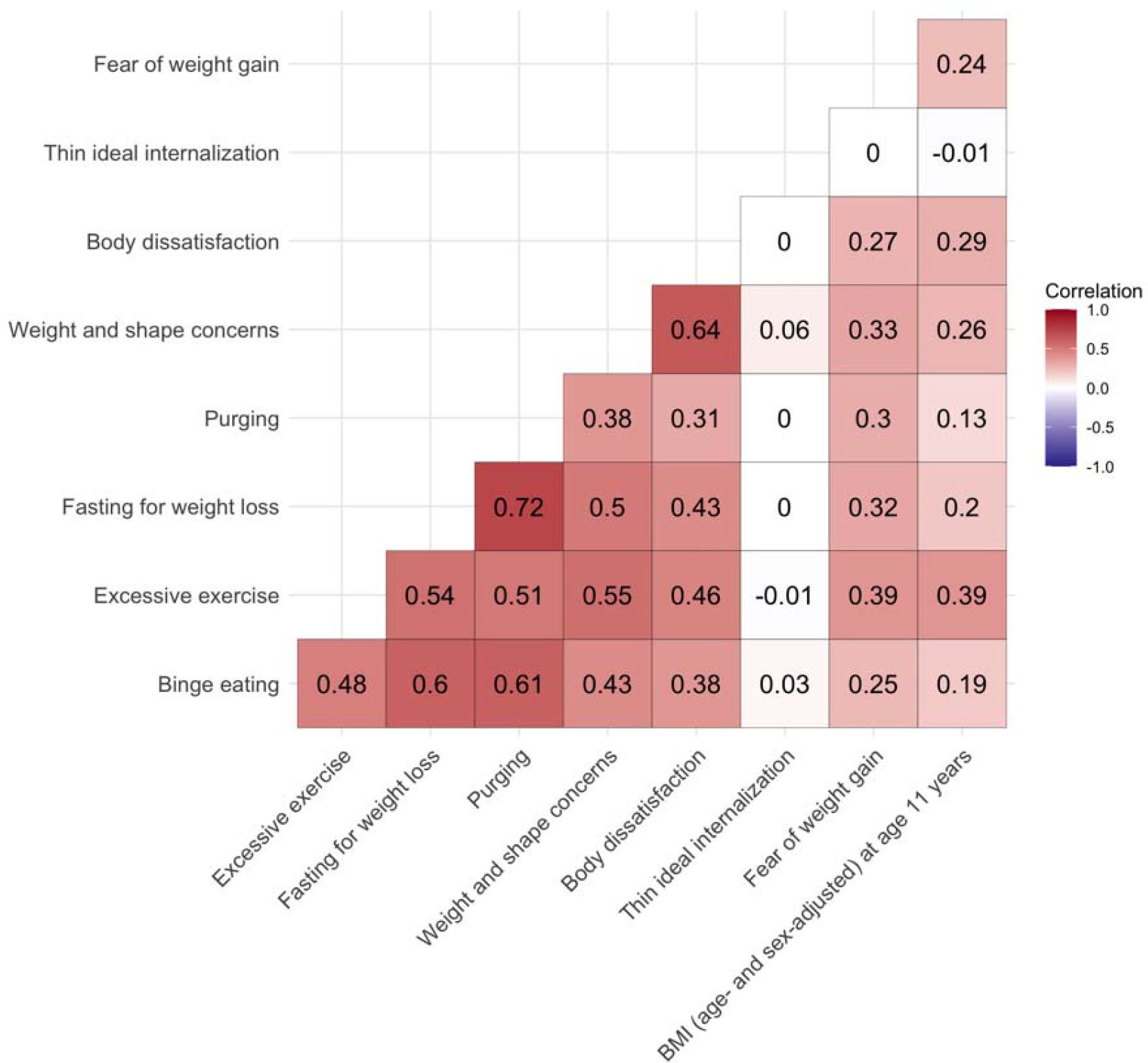
Heterogenous correlation matrix of the investigated eating disorder (ED) symptoms and body mass index (BMI) in the Avon Longitudinal Study of Parents and Children (ALSPAC) (Boyd et al., 2013; Fraser et al., 2013; Golding, 2004) using the R package ‘polycor’ (version 0.7-10). Sample sizes per outcome were binge eating (*n* cases = 840, *n* controls = 2151), excessive exercise (*n* cases = 2,332, *n* controls = 2,893), fasting for weight loss (*n* cases = 777, *n* controls = 2,678), fear of weight gain (*n* = 6,013, mean = 1.62, SD = 1.24), purging (*n* cases = 398, *n* controls = 2,962), body dissatisfaction (*n* = 4,624, mean = 21.85, SD = 7.75), thin ideal internalization (*n* = 4,495, mean = 15.33, SD = 2.69), weight and shape concerns (*n* = 4,622, mean = 5.34, SD = 1.85), zBMI at age 11 years (*n* = 4,219, mean = 0.27, SD = 1.16). SD = standard deviation.

**Table 1:** Descriptive statistics of the binary eating disorder (ED) symptoms outcomes of the participants in the Avon Longitudinal Study of Parents and Children^a^

| Eating disorder symptom | Age phenotype measured | Total N | Cases N (%) | Cases (% female) | Controls N | Controls (% female) |
| --- | --- | --- | --- | --- | --- | --- |
| Binge eating | 14,16,18 | 2,991 | 840 (28.08) | 78.81 | 2,151 | 54.3 |
| Excessive exercise | 14,16,18 | 5,225 | 2,332 (44.63) | 72.14 | 2,893 | 42.62 |
| Fasting for weight loss | 14,16,18 | 3,455 | 777 (22.49) | 87.52 | 2,678 | 53.02 |
| Purging | 14,16,18 | 3,360 | 398 (11.85) | 88.94 | 2,962 | 56.25 |

<sup>a</sup> Full description of the Avon Longitudinal Study of Parents and Children is described elsewhere (Boyd et al., 2013; Fraser et al., 2013; Golding, 2004). The descriptive depicted in this table are on individuals for which genotype information was available.

**Table 2:**
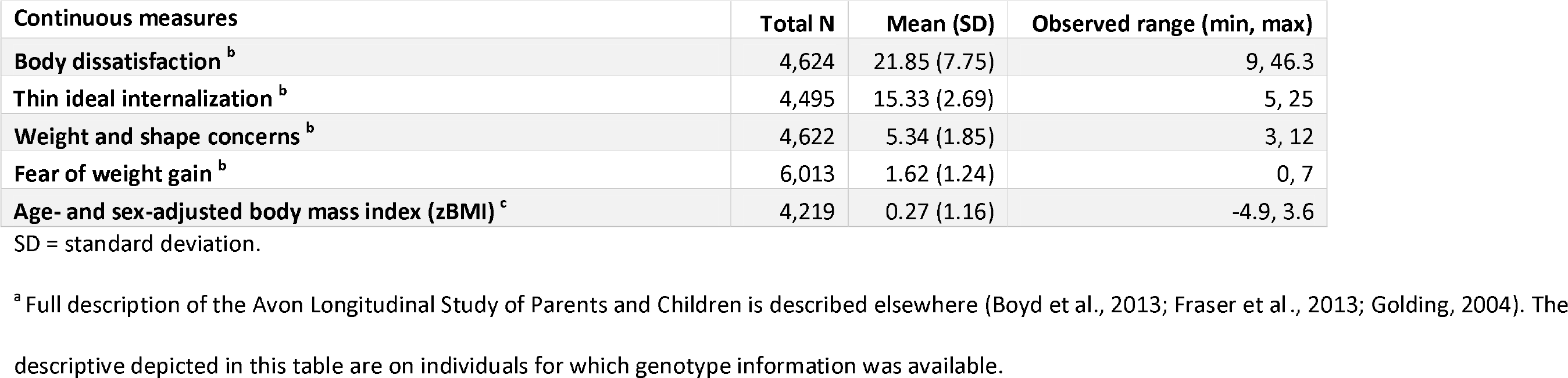
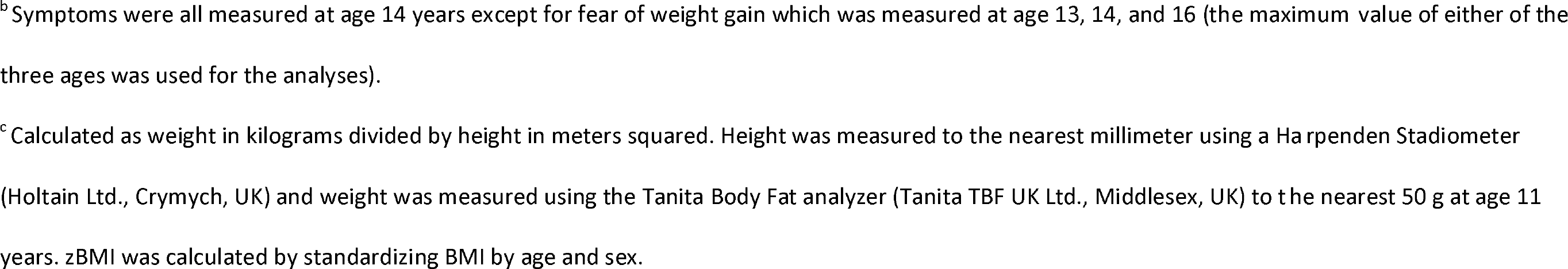
Descriptive statistics of the continuous eating disorder (ED) symptoms outcomes and body mass of the participants in the Avon Longitudinal Study of Parents and Children^a^

| Continuous measures | Total N | Mean (SD) | Observed range (min, max) |
| --- | --- | --- | --- |
| Body dissatisfaction <sup>b</sup> | 4,624 | 21.85 (7.75) | 9, 46.3 |
| Thin ideal internalization <sup>b</sup> | 4,495 | 15.33 (2.69) | 5, 25 |
| Weight and shape concerns <sup>b</sup> | 4,622 | 5.34 (1.85) | 3, 12 |
| Fear of weight gain <sup>b</sup> | 6,013 | 1.62 (1.24) | 0, 7 |
| Age- and sex-adjusted body mass index (zBMI) <sup>c</sup> | 4,219 | 0.27 (1.16) | -4.9, 3.6 |

SD = standard deviation.
<sup>a</sup> Full description of the Avon Longitudinal Study of Parents and Children is described elsewhere (Boyd et al., 2013; Fraser et al., 2013; Golding, 2004). The descriptive depicted in this table are on individuals for which genotype information was available.
<sup>b</sup> Symptoms were all measured at age 14 years except for fear of weight gain which was measured at age 13, 14, and 16 (the maximum value of either of the three ages was used for the analyses).
<sup>c</sup> Calculated as weight in kilograms divided by height in meters squared. Height was measured to the nearest millimeter using a Harpenden Stadiometer (Holtain Ltd., Crymych, UK) and weight was measured using the Tanita Body Fat analyzer (Tanita TBF UK Ltd., Middlesex, UK) to the nearest 50 g at age 11 years. zBMI was calculated by standardizing BMI by age and sex.

#### Polygenic scores associated with ED symptoms

After correcting for multiple testing using the false discovery rate adjustment, eight PGS were significantly associated with ED symptoms in ALSPAC. Reported are odds ratios per one standard deviation increase in the PGS for the binary ED symptoms (i.e., binge eating, excessive exercise, fasting for weight loss, and purging) and betas per one standard deviation increase in the PGS for the continuous ED symptoms (i.e., body dissatisfaction, thin ideal internalization, weight and shape concerns, and fear of weight gain); **Table 3, Figure 2-3, and Supplementary Tables S2-3.**

**Figure 2:**
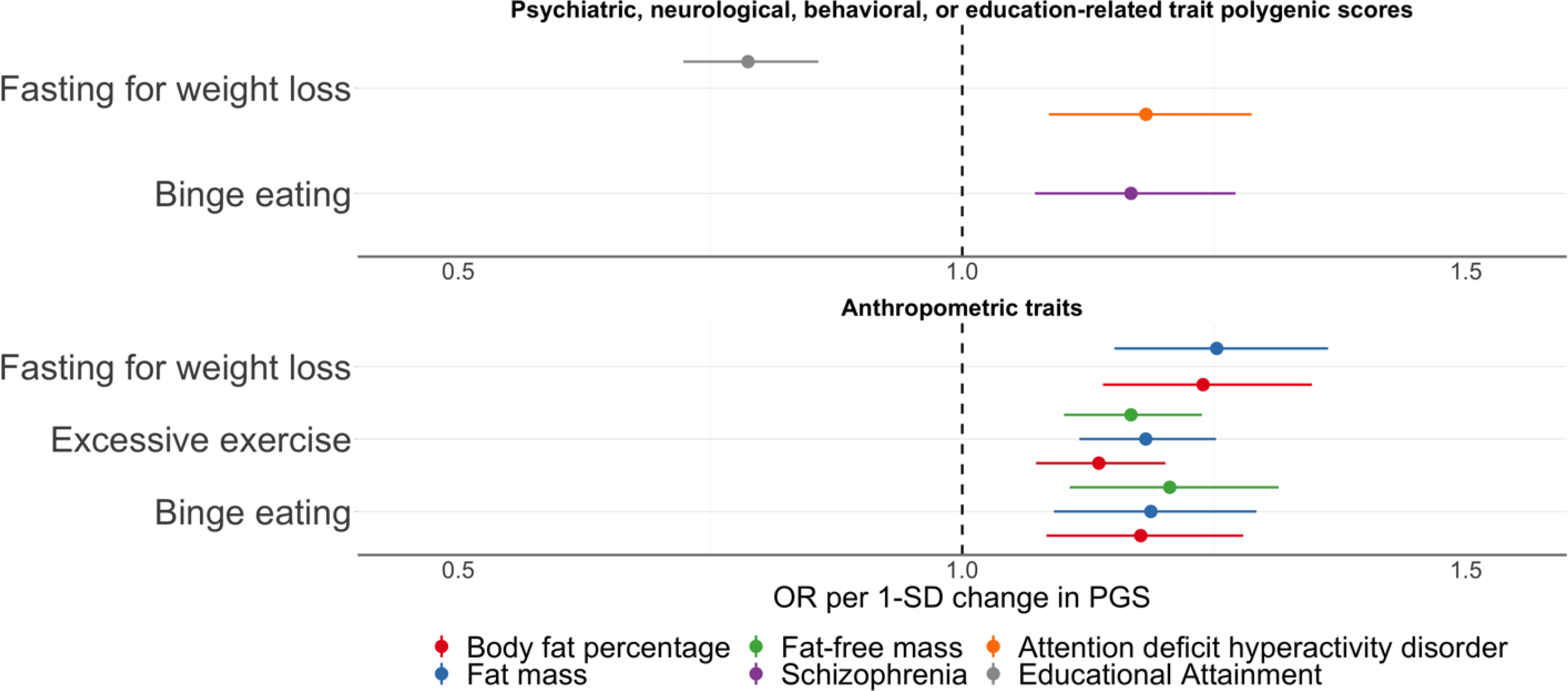
Polygenic scores associated with binge eating, excessive exercise, and fasting for weight loss in the Avon Longitudinal Study of Parents and Children. Sample sizes per outcome were binge eating (*n* cases = 840, *n* controls = 2151), excessive exercise (*n* cases = 2,332, *n* controls = 2,893) fasting for weight loss (*n* cases = 777, *n* controls = 2,678). Polygenic scores were calculated using PRSice v2 (Choi & O’Reilly, 2019). The optimal *p* value thr shold to generate the polygenic score was obtained by calculating polygenic scores across multiple thresholds and permuting case-control status at each threshold 10,000 times. The polygenic score explaining the largest trait variance was used in logistic regressions including the first six ancestry principal componentsand sex as covariates. Shown here are only the associations between the polygenic scores and the binary eating disorder symptoms that remained significant after correction for multiple testing via calculation of false discovery rate-adjusted *Q* values (refer to **Tables S2-S3** for a complete overview of the results). The dots represent the point estimates of the odd ratios for an increase of one standard deviation in the polygenic score and the lines represent the 95% confidence interval of the point estimate.

**Figure 3:**
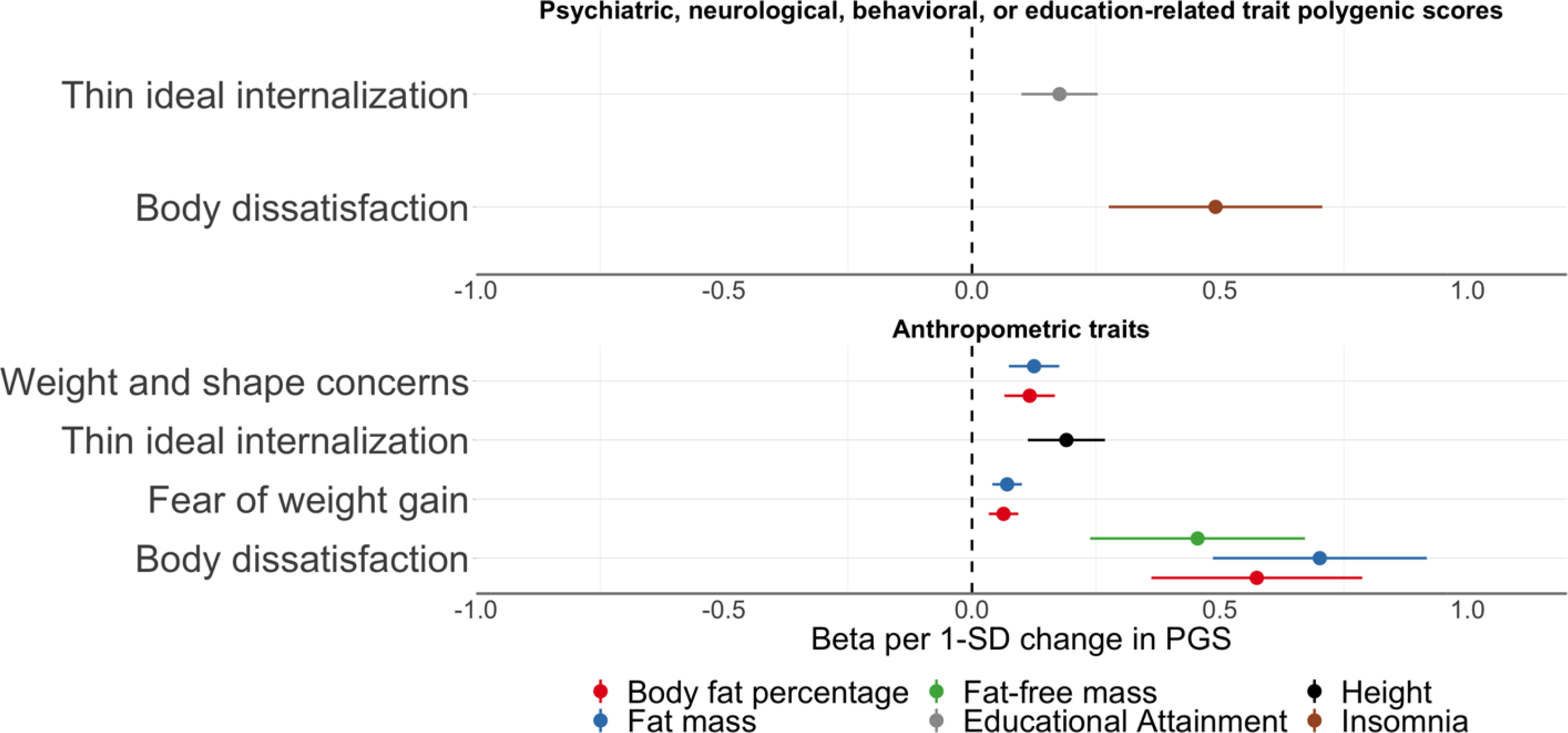
Polygenic scores associated with body dissatisfaction, fear of weight gain, thin ideal internalization, and weight and shape concerns in the Avon Longitudinal Study of Parents and Children. Sample sizes per outcome were body dissatisfaction (*n* =4,624, *mean* = 21.85, *SD* = 7.75), fear of w ight gain (*n* =6,013, *mean* = 1.62, *SD* = 1.24), thin ideal internalization (*n* = 4,495, *mean* = 15.33, *SD* = 2.69), and weight and shape concerns (*n* =4,622, *mean* = 5.34, *SD* = 1.85). Polygenic scores were calculated using PRSice v2. The optimal *p* value threshold to generate the polygenic score was obtained by calculating polygenic scores across multiple thresholds and permuting case-control status at each threshold 10,000 times. The polygenic score explaining the largest trait variance was used in linear regressions including the first six ancestry principal components and sex as covariates. Shown here are only the associations between the polygenic scores and the continuous eating disorder symptoms that remained significant after correction for multiple testing via calculation of false discovery rate-adjusted *Q* values (refer to **Tables S2-S3** for a complete overview of the results). The dots represent the point estimates of the β for an increase of one standard deviation in the PGS and the lines represent the 95% confidence interval of the point estimate.

**Table 3:**
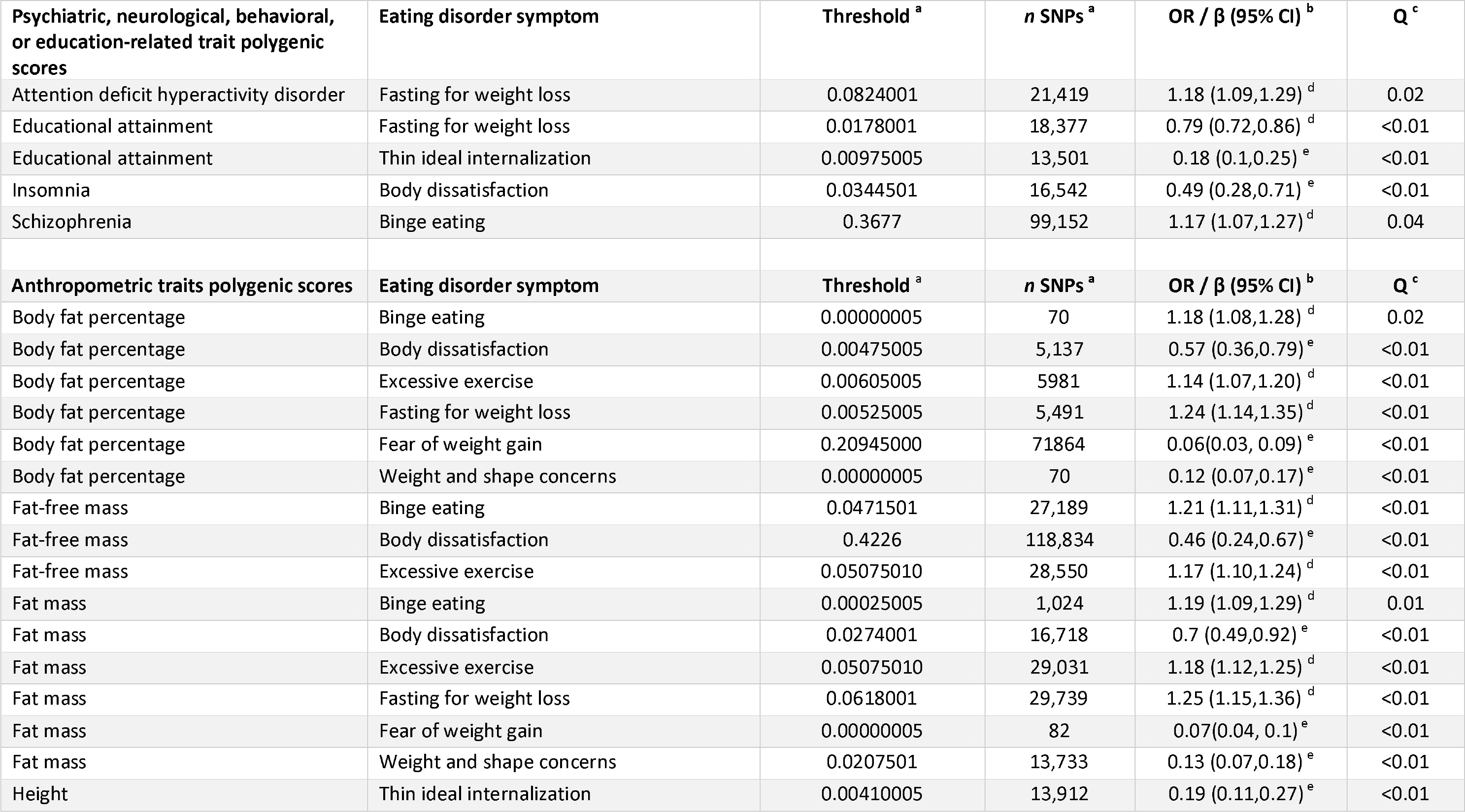
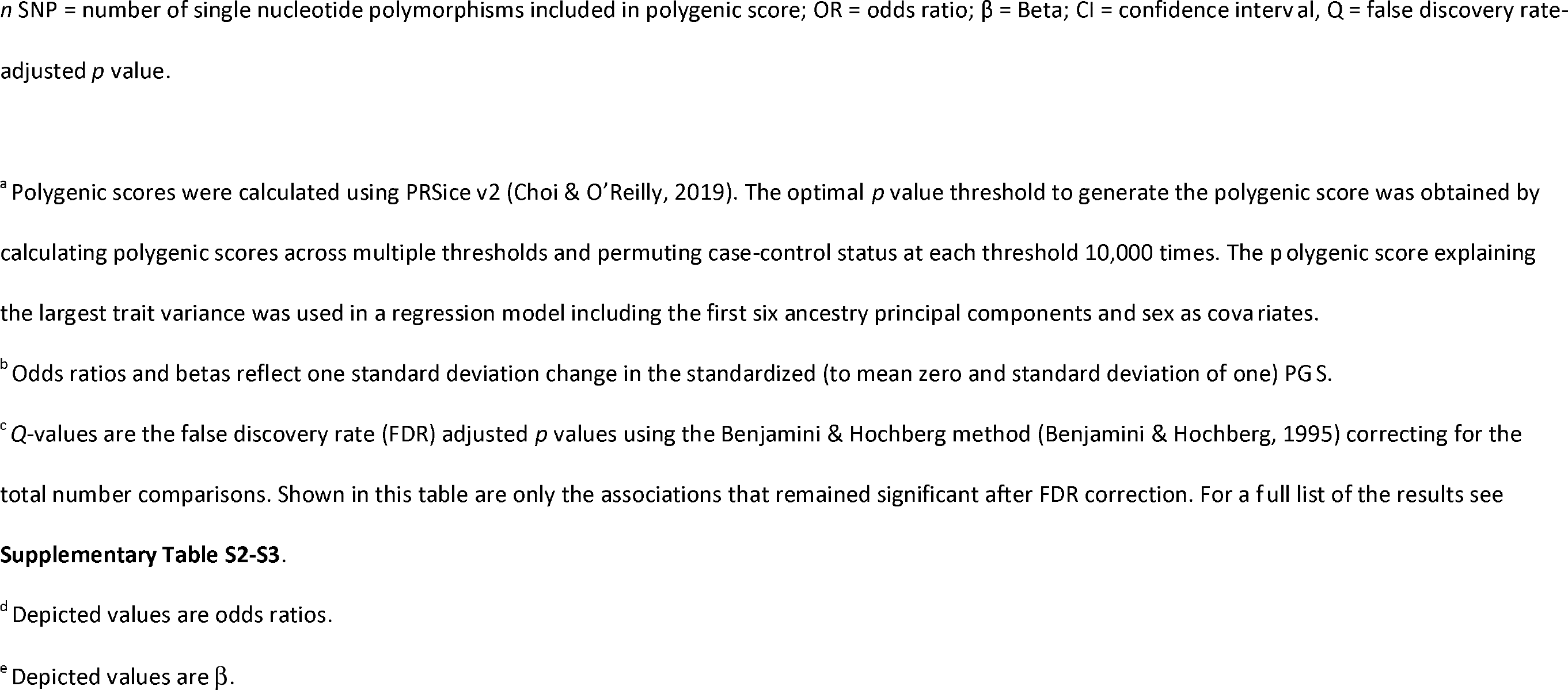
Associations of psychiatric, metabolic, and anthropometric polygenic scores (PGSs) with eating disorder (ED) symptoms

#### Association with psychiatric, neurological, behavioral, or education-related traits PGSs

Four PGSs reflecting polygenic liability to psychiatric disorders or an education-related trait showed significant associations with ED symptoms (**Table 3, Figure 2-3**). The schizophrenia PGS was positively associated with binge eating (OR per standard deviation in PGS = 1.17, 95% confidence interval [CI]: 1.07, 1.27; *Q* = 0.04). Higher insomnia PGS was positively associated with body dissatisfaction (β = 0.49, 95% CI: 0.28, 0.71; *Q* < 0.01). Individuals with higher educational attainment PGS were less likely to engage in fasting behavior (OR = 0.79, 95% CI: 0.72, 0.86; *Q* < 0.01), and had on average higher scores on thin ideal internalization compared to individuals with lower PGS on educational attainment (β = 0.18, 95% CI: 0.10, 0.25; *Q* < 0.01). Additionally, individuals with a standard deviation higher ADHD PGS were more likely to engage in fasting behavior (OR = 1.18, 95% CI: 1.09, 1.29; *Q* = 0.03). We found no evidence of association for the psychiatric, neurological, behavioral, or education-related trait PGSs with excessive exercise and weight and shape concerns.

#### Association with metabolic trait PGSs

None of the PGSs indexing metabolic traits were associated with any ED symptoms, after correcting for multiple comparisons (**Supplementary Tables S2-3**).

#### Association with anthropometric trait PGSs

Several PGSs of anthropometric traits were positively associated with ED symptoms (**Table 3, Figure 2-3, and Supplementary Tables S2-3**). For instance, the height PGS was positively associated with thin ideal internalization (β= 0.19, 95% CI: 0.11, 0.27; *Q* < 0.01), whereas the fat-free mass PGS was positively associated with binge eating (OR = 1.21, 95% CI: 1.11, 1.31; *Q* < 0.01), excessive exercise (OR = 1.17, 95% CI: 1.10, 1.24; *Q* < 0.01), and body dissatisfaction (β = 0.46, 95% CI: 0.24, 0.67; *Q* < 0.01). A similar pattern was observed for the fat mass PGS which was positively associated with binge eating (OR = 1.19, 95% CI: 1.09, 1.29; *Q* = 0.01), excessive exercise (OR = 1.18, 95% CI: 1.12, 1.25; *Q* < 0.01), fasting (OR = 1.25, 95% CI: 1.15, 1.36; *Q* < 0.01), body dissatisfaction (β = 0.70, 95% CI: 0.49, 0.92; *Q* < 0.01), and weight and shape concerns (β = 0.13, 95% CI: 0.07, 0.18; *Q* < 0.01). The body fat percentage PGS showed the same pattern as the fat mass PGS; that is, a positive association with binge eating (OR = 1.18, 95% CI: 1.08, 1.28; *Q* = 0.02), excessive exercise (OR = 1.14, 95% CI: 1.07, 1.20; *Q* < 0.01), fasting (OR = 1.24, 95% CI: 1.14, 1.35; *Q* < 0.01), body dissatisfaction (β = 0.57, 95% CI: 0.36, 0.79; *Q* < 0.01), weight and shape concerns (β = 0.12, 95% CI: 0.07, 0.17; *Q* < 0.01). The similarity of the results is likely due to either phenotypic correlations (e.g., fasting for weight loss and binge eating *r* = 0.60; **Figure 1**), correlation between the PGSs (fat mass and body fat percentage *r* = 0.89; **Supplemental Figure S1**), or a combination of both.

#### Exploratory causal mediation analyses

Prior to inclusion in the mediation analyses, we tested whether the ADHD, educational attainment, insomnia, and the schizophrenia PGS were associated with BMI at age 11 years (**Supplementary Table S4**); only the ADHD (β = 0.13, 95% CI: 0.06, 0.20; *Q* < 0.01) and the educational attainment PGS (β = -0.19, 95% CI: - 0.26, -0.12; *Q* < 0.01) were significantly associated with BMI at age 11 years and were therefore included in the causal mediation analyses. Childhood BMI significantly mediated the association between the ADHD PGS and fasting for weight loss (β average causal mediation effect = 0.004, 95% CI: 0.002, 0.007; *p* < 0.01); one SD increase in the ADHD PGS corresponded with a higher BMI at age 11 years, which in turn increased the probability of endorsing fasting for weight loss. Furthermore, we found that BMI significantly mediated the association between the educational attainment PGS and fasting for weight loss (β average causal mediation effect = -0.004, 95% CI: -0.006, -0.002; *p* < 0.01; **Table 4**); that is, a one standard deviation increase in the educational attainment PGS corresponded with a decrease in BMI at age 11 years, which in turn led to a decreased probability of endorsing fasting for weight loss during adolescence. However, we did not observe a significant mediation effect by BMI in the association between the educational attainment PGS and thin ideal internalization (β average causal mediation effect = -0.002, 95% CI: -0.005, 0.001; *p* = 0.27).

**Table 4:**
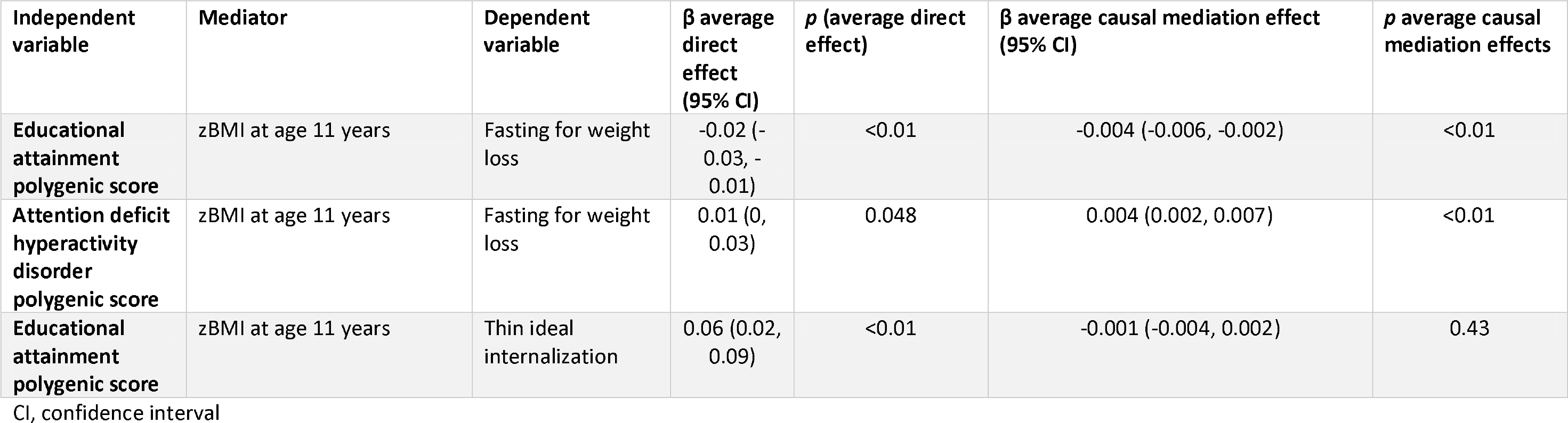
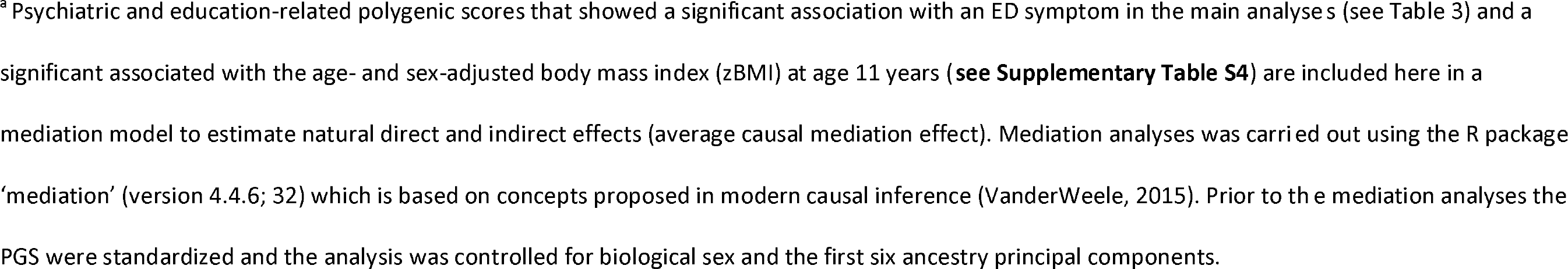
Causal mediation analysis of the attention deficit hyperactivity disorder and the educational attainment polygenic score scores and the eating disorder (ED) symptoms fasting for weight loss and thin ideal internalization with the age- and sex-adjusted body mass index (zBMI) at age 11 years as mediator.*^a^* *P*-values for mediation were generated with bootstrapping methods.

## Discussion

We report molecular genetic evidence that genetics of psychiatric disorders and anthropometric traits, but not metabolic traits, as indexed by PGSs are associated with ED symptoms and traits present in a general population cohort in adolescence. Notably, several ED symptoms (i.e., binge eating, fasting for weight loss, and body dissatisfaction) in this study were significantly associated with both psychiatric and anthropometric PGSs emphasizing the genetic complexity of these traits and both psychiatric and anthropometric etiological contributions. Importantly, contrary to our hypotheses, and unlike previously published results for AN, ED symptoms were not associated with the selected metabolic PGSs (i.e., type 2 diabetes, insulin resistance, fasting insulin, and high-density lipoprotein [HDL] cholesterol). This pattern suggests that ED symptoms in the population and threshold EDs may be partially etiologically related (i.e., psychiatric and anthropometric origins), but that metabolic genetic factors may differentiate between symptoms and threshold EDs.

Our results highlight novel associations between educational attainment and ED symptoms. An increase in educational attainment PGS was associated with lower likelihood of endorsing fasting for weight loss and higher levels of thin ideal internalization (the extent to which a person agrees with socially defined ideals of attractiveness, such as the notion that thin individuals are attractive). In previous work, we demonstrated that fasting for weight loss was positively associated with BMI (Abdulkadir et al., 2020) and given reports of a causal relationship between higher education and decreased fat mass (Hübel et al., 2019), it is not surprising that the educational attainment PGS was negatively associated with fasting for weight loss. Further exploration of the association between the educational attainment PGS and fasting for weight loss revealed that this association is mediated through BMI; an increase in educational attainment PGS leads to a decrease in childhood BMI which in turn leads to a lower probability of endorsing fasting for weight loss during adolescence. Furthermore, the positive association between higher educational attainment PGS and thin ideal internalization is consistent with several studies that reported positive genetic correlation (*r*_g_ = 0.25) between educational attainment and AN (Duncan et al., 2017; Watson et al., 2019); our measure of thin ideal internalization captures body image disturbances that are considered a core component of AN. Interestingly, in contrast to our finding for fasting for weight loss, the association between the educational attainment PGS and thin ideal internalization was not mediated through BMI. Taken together, our findings suggest that thin ideal internalization as measured in the general population could share the same genetic risk as AN in part through alleles that influence educational attainment independent of BMI.

Another novel finding to emerge from our study is the positive association between the insomnia PGS and body dissatisfaction; a finding that aligns with previous reports (Akram, 2017; Gupta, Gupta, & Knapp, 2015) of positive associations between self-reported insomnia symptoms and dissatisfaction with cutaneous body image (mental perception of the appearance of the skin, hair, and nails). We sought to further understand the association between the insomnia PGS and body dissatisfaction by testing whether BMI is a possible mediator; body dissatisfaction was associated with BMI in this sample (Abdulkadir et al., 2020) and could therefore operate as a mediator. Counter to our expectation, we found no association between the insomnia PGS and childhood BMI suggesting that childhood BMI is likely not on the causal pathway between the insomnia PGS and body dissatisfaction in adolescence.

Returning to the absence of a significant association between metabolic PGSs and ED symptoms, alternative interpretations exist. First, the finding might be true, and the absence of metabolic effects could represent a point of divergence between ED symptoms and threshold disorders. Alternatively, the majority of ED symptoms studied here (apart from thin ideal internalization and fear of weight gain) are more closely related to bulimic type disorders, and no GWAS of binge-type eating disorders have yet been conducted. The metabolic PGSs that we selected were based on significant correlations observed in an AN GWAS. Emerging bulimia and binge-eating GWAS may implicate different metabolic factors that were not captured here. If the result holds, genetically influenced metabolic vulnerability may represent the “tipping point” between subthreshold symptoms and threshold disorders. This notion aligns with findings from studies that suggest that AN might be more distinctly genetically demarcated from ED symptoms in the general population than other EDs (Adams, Reay, Geaghan, & Cairns, 2020; Dinkler et al., 2019).

Purging stood out in our analyses as it was not associated with any of the twenty-five PGSs. It is likely that our analyses were underpowered for purging as this trait was the least frequently endorsed in our study population (*N* case = 398).

Our findings must be interpreted in light of the following limitations. The ED symptoms were self-reported, and the questions pertained to the previous year, which may have resulted in recall bias. Ideally, clinician-assessed measures are preferred, but this is not feasible for large cohort studies, such as ALSPAC. Furthermore, the sample only included white British participants and therefore the results of this study are not generalizable to other populations. Like many other longitudinal studies, in the ALSPAC study, participants tend to drop out as time goes on leading to missing data. However, we would like to emphasize that the longitudinal nature of the study allowed us to define phenotypes across different timepoints, which in part helped in mitigating missingness. Furthermore, there is evidence that indicates that higher PGSs on smoking initiation, schizophrenia, and depression in ALSPAC participants are associated with higher dropout rates from the study (Taylor et al., 2018); therefore, any bias introduced by higher dropout likely attenuated associations toward the null.

The associations between ED symptoms, that are widely prevalent in the community, and psychiatric and anthropometric PGSs highlight that these symptoms are polygenic and may share a genetic architecture with AN but yet could differ from AN in one important component, namely metabolic disturbances. It is critical to understand why some individuals who endorse ED symptoms progress to develop threshold AN whereas others do not; metabolic disturbances could be the catalyst that puts some individuals on a developmental trajectory leading to AN. Future studies are needed to understand when during development these metabolic disturbances manifest and how they may affect developmental trajectory of AN. A better understanding of this process could bring us closer to improved therapeutics for AN.

In conclusion, our results suggest that ED symptoms as present in the general population are genetically associated with psychiatric and anthropometric traits but not with metabolic traits. Our findings serve as a base for future translational research aimed at understanding the potential shared biological mechanisms among ED symptoms, psychiatric traits, and anthropometric traits, and differences in risk for ED symptoms vs. threshold EDs, as implied by the findings of this study.

## Data Availability

Please note that the study website contains details of all the data that is available through a fully searchable data dictionary and variable search tool" and reference the following webpage: http://www.bristol.ac.uk/alspac/researchers/our-data/.

## Acknowledgements

We are extremely grateful to all the families who took part in this study, the midwives for their help in recruiting them, and the whole ALSPAC team, which includes interviewers, computer and laboratory technicians, clerical workers, research scientists, volunteers, managers, receptionists and nurses.

## Author Contribution

MA, CH, MH analyzed the data. MA, CH, MH drafted the manuscript. NM, CMB, RFL, and GB supervised the work. All authors substantially contributed to the conception and interpretation of the work, revised the manuscript for important intellectual content and approved the final version. All authors agree to be accountable for all aspects of this work.

## References

Abdulkadir, M., Herle, M., De Stavola, B. L., Hübel, C., Santos Ferreira, D. L., Loos, R. J. F., … Micali, N. (2020). Polygenic Score for Body Mass Index Is Associated with Disordered Eating in a General Population Cohort. Journal of Clinical Medicine, 9(4), 1187. https://doi.org/10.3390/jcm9041187

Adams, D. M., Reay, W. R., Geaghan, M. P., & Cairns, M. J. (2020). Investigation of glycaemic traits in psychiatric disorders using Mendelian randomisation revealed a causal relationship with anorexia nervosa. Neuropsychopharmacology, (August), 1–10. https://doi.org/10.1038/s41386-020-00847-w

Akram, U. (2017). Sleep associated monitoring on awakening mediates the relationship between cutaneous body image dissatisfaction and insomnia symptoms. Sleep Science, 10(2), 92–95. https://doi.org/10.5935/1984-0063.20170017

Arnold, P. D., Askland, K. D., Barlassina, C., Bellodi, L., Bienvenu, O. J., Black, D., … Zai, G. (2018). Revealing the complex genetic architecture of obsessive–compulsive disorder using meta-analysis. Molecular Psychiatry, 23(5), 1181–1188. https://doi.org/10.1038/mp.2017.154

Benjamini, Y., & Hochberg, Y. (1995). Controlling the False Discovery Rate : a Practical and Powerful Approach to Multiple Testing. Journal of the Royal Statistical Society. Series B (Methodological), 57(1), 289–300.

Benson, R., von Hippel, P. T., & Lynch, J. L. (2018). Does more education cause lower BMI, or do lower-BMI individuals become more educated? Evidence from the National Longitudinal Survey of Youth 1979. Social Science & Medicine, 211, 370–377. https://doi.org/10.1016/j.socscimed.2017.03.042

Boraska, V., Davis, O. S. P., Cherkas, L. F., Helder, S. G., Harris, J., Krug, I., … Zeggini, E. (2012). Genome-wide association analysis of eating disorder-related symptoms, behaviors, and personality traits. American Journal of Medical Genetics Part B: Neuropsychiatric Genetics, 159B(7), 803–811. https://doi.org/10.1002/ajmg.b.32087

Boyd, A., Golding, J., Macleod, J., Lawlor, D. A., Fraser, A., Henderson, J., … Smith, G. D. (2013). Cohort profile: The ’Children of the 90s’-The index offspring of the avon longitudinal study of parents and children. International Journal of Epidemiology, 42(1), 111–127. https://doi.org/10.1093/ije/dys064

Boyd, A., Thomas, R., Hansell, A. L., Gulliver, J., Hicks, L. M., Griggs, R., … Macleod, J. (2019). Data Resource Profile: The ALSPAC birth cohort as a platform to study the relationship of environment and health and social factors. International Journal of Epidemiology, 48(4), 1038–1039k. https://doi.org/10.1093/ije/dyz063

Calzo, J. P., Austin, S. B., & Micali, N. (2018). Sexual orientation disparities in eating disorder symptoms among adolescent boys and girls in the UK. European Child and Adolescent Psychiatry, 27(11), 1–8. https://doi.org/10.1007/s00787-018-1145-9

Chamay-Weber, C., Narring, F., & Michaud, P.-A. (2005). Partial eating disorders among adolescents: A review. Journal of Adolescent Health, 37(5), 416–426. https://doi.org/10.1016/j.jadohealth.2004.09.014

Choi, S. W., & O’Reilly, P. F. (2019). PRSice-2: Polygenic Risk Score software for biobank-scale data. GigaScience, 8(7), 1–6. https://doi.org/10.1093/gigascience/giz082

Cole, T. J., Bellizzi, M., Flegal, K., & Dietz, W. (2000). Establishing a standard definition for child overweight and obesity worldwide: international survey. BMJ, 320(7244), 1240– 1240. https://doi.org/10.1136/bmj.320.7244.1240

Crow, S., Eisenberg, M. E., Story, M., & Neumark-Sztainer, D. (2008). Are body dissatisfaction, eating disturbance, and body mass index predictors of suicidal behavior in adolescents? A longitudinal study. Journal of Consulting and Clinical Psychology, 76(5), 887–892. https://doi.org/10.1037/a0012783

Demontis, D., Walters, R. K., Martin, J., Mattheisen, M., Als, T. D., Agerbo, E., … Neale, B. M. (2019). Discovery of the first genome-wide significant risk loci for attention deficit/hyperactivity disorder. Nature Genetics, 51(1), 63–75. https://doi.org/10.1038/s41588-018-0269-7

Dinkler, L., Taylor, M. J., Råstam, M., Hadjikhani, N., Bulik, C. M., Lichtenstein, P., … Lundström, S. (2019). Association of etiological factors across the extreme end and continuous variation in disordered eating in female Swedish twins. Psychological Medicine. https://doi.org/10.1017/S0033291719003672

Duncan, L., Ratanatharathorn, A., Aiello, A. E., Almli, L. M., Amstadter, A. B., Ashley-Koch, A. E., … Koenen, K. C. (2018). Largest GWAS of PTSD (N=20 070) yields genetic overlap with schizophrenia and sex differences in heritability. Molecular Psychiatry, 23(3), 666– 673. https://doi.org/10.1038/mp.2017.77

Duncan, L., Yilmaz, Z., Gaspar, H., Walters, R., Goldstein, J., Anttila, V., … Bulik, C. M. (2017). Significant Locus and Metabolic Genetic Correlations Revealed in Genome-Wide Association Study of Anorexia Nervosa. American Journal of Psychiatry, 174(9), 850– 858. https://doi.org/10.1176/appi.ajp.2017.16121402

Dupuis, J., Langenberg, C., Prokopenko, I., Saxena, R., Soranzo, N., Jackson, A. U., … Wilson, J. F. (2010). New genetic loci implicated in fasting glucose homeostasis and their impact on type 2 diabetes risk. Nature Genetics, 42(2), 105–116. https://doi.org/10.1038/ng.520

Field, A. E., Taylor, C. B., Celio, A., & Colditz, G. A. (2004). Comparison of self-report to interview assessment of bulimic behaviors among preadolescent and adolescent girls and boys. International Journal of Eating Disorders, 35(1), 86–92. https://doi.org/10.1002/eat.10220

Fraser, A., Macdonald-wallis, C., Tilling, K., Boyd, A., Golding, J., Davey smith, G., … Lawlor, D. A. (2013). Cohort profile: The avon longitudinal study of parents and children: ALSPAC mothers cohort. International Journal of Epidemiology, 42(1), 97–110. https://doi.org/10.1093/ije/dys066

Golding, J. (2004). The Avon Longitudinal Study of Parents and Children (ALSPAC)--study design and collaborative opportunities. *European Journal of Endocrinology*, U119– U123. https://doi.org/10.1530/eje.0.151u119

Golding, Pembrey, Jones, & The Alspac Study Team. (2001). ALSPAC-The Avon Longitudinal Study of Parents and Children. Paediatric and Perinatal Epidemiology, 15(1), 74–87. https://doi.org/10.1046/j.1365-3016.2001.00325.x

Gormley, P., Anttila, V., Winsvold, B. S., Palta, P., Esko, T., Pers, T. H., … Palotie, A. (2016). Meta-analysis of 375,000 individuals identifies 38 susceptibility loci for migraine. Nature Genetics, 48(8), 856–866. https://doi.org/10.1038/ng.3598

Grove, J., Ripke, S., Als, T. D., Mattheisen, M., Walters, R. K., Won, H., … Børglum, A. D. (2019). Identification of common genetic risk variants for autism spectrum disorder. Nature Genetics. https://doi.org/10.1038/s41588-019-0344-8

Gupta, M. A., Gupta, A. K., & Knapp, K. (2015). Dissatisfaction with cutaneous body image is directly correlated with insomnia severity: A prospective study in a non-clinical sample. Journal of Dermatological Treatment, 26(2), 193–197. https://doi.org/10.3109/09546634.2014.883060

Hayes, J. F., Fitzsimmons-Craft, E. E., Karam, A. M., Jakubiak, J., Brown, M. L., & Wilfley, D. E. (2018). Disordered Eating Attitudes and Behaviors in Youth with Overweight and Obesity: Implications for Treatment. Current Obesity Reports, 7(3), 235–246. https://doi.org/10.1007/s13679-018-0316-9

Hübel, C., Gaspar, H. A., Coleman, J. R. I., Hanscombe, K. B., Purves, K., Prokopenko, I., … Breen, G. (2019). Genetic correlations of psychiatric traits with body composition and glycemic traits are sex- and age-dependent. Nature Communications, 10(1), 5765. https://doi.org/10.1038/s41467-019-13544-0

Hübel, C., Gaspar, H. A., Coleman, J. R. I. I., Finucane, H., Purves, K. L., Hanscombe, K. B., … Breen, G. (2019). Genomics of body fat percentage may contribute to sex bias in anorexia nervosa. American Journal of Medical Genetics, Part B: Neuropsychiatric Genetics, 180(6), 428–438. https://doi.org/10.1002/ajmg.b.32709

Jaacks, L. M., Vandevijvere, S., Pan, A., McGowan, C. J., Wallace, C., Imamura, F., … Ezzati, M. (2019). The obesity transition: stages of the global epidemic. The Lancet. Diabetes & Endocrinology, 7(3), 231–240. https://doi.org/10.1016/S2213-8587(19)30026-9

Jansen, P. R., Watanabe, K., Stringer, S., Skene, N., Bryois, J., Hammerschlag, A. R., … Posthuma, D. (2019). Genome-wide analysis of insomnia in 1,331,010 individuals identifies new risk loci and functional pathways. Nature Genetics, 51(3), 394–403. https://doi.org/10.1038/s41588-018-0333-3

Kann, L., Warren, C. W., Harris, W. A., Collins, J. L., Williams, B. I., Ross, J. G., & Kolbe, L. J. (1996). Youth Risk Behavior Surveillance-United States, 1995. Journal of School Health, 66(10), 365–377. https://doi.org/10.1111/j.1746-1561.1996.tb03394.x

Lee, J. J., Wedow, R., Okbay, A., Kong, E., Maghzian, O., Zacher, M., … Cesarini, D. (2018). Gene discovery and polygenic prediction from a genome-wide association study of educational attainment in 1.1 million individuals. Nature Genetics, 50(8), 1112–1121.https://doi.org/10.1038/s41588-018-0147-3

McCrea, R. L., Berger, Y. G., & King, M. B. (2012). Body mass index and common mental disorders: exploring the shape of the association and its moderation by age, gender and education. International Journal of Obesity, 36(3), 414–421. https://doi.org/10.1038/ijo.2011.65

Micali, N., De Stavola, B., Ploubidis, G., Simonoff, E., Treasure, J., & Field, A. E. (2015). Adolescent eating disorder behaviours and cognitions: Gender-specific effects of child, maternal and family risk factors. British Journal of Psychiatry, 207(4), 320–327. https://doi.org/10.1192/bjp.bp.114.152371

Micali, N., Horton, N. J., Crosby, R. D., Swanson, S. A., Sonneville, K. R., Solmi, F., … Field, A. E. (2017). Eating disorder behaviours amongst adolescents: investigating classification, persistence and prospective associations with adverse outcomes using latent class models. European Child & Adolescent Psychiatry, 26(2), 231–240. https://doi.org/10.1007/s00787-016-0877-7

Nagata, J. M., Braudt, D. B., Domingue, B. W., Bibbins-Domingo, K., Garber, A. K., Griffiths, S., & Murray, S. B. (2019). Genetic risk, body mass index, and weight control behaviors: Unlocking the triad. International Journal of Eating Disorders, 52(7), 825–833. https://doi.org/10.1002/eat.23083

Northstone, K., Lewcock, M., Groom, A., Boyd, A., Macleod, J., Timpson, N., & Wells, N. (2019). The Avon Longitudinal Study of Parents and Children (ALSPAC): an update on the enrolled sample of index children in 2019. Wellcome Open Research, 4, 51. https://doi.org/10.12688/wellcomeopenres.15132.1

Pasman, J. A., Verweij, K. J. H., Gerring, Z., Stringer, S., Sanchez-Roige, S., Treur, J. L., … Vink, J. M. (2018). GWAS of lifetime cannabis use reveals new risk loci, genetic overlap with psychiatric traits, and a causal effect of schizophrenia liability. Nature Neuroscience, 21(9), 1161–1170. https://doi.org/10.1038/s41593-018-0206-1

Purcell, S. M., Wray, N. R., Stone, J. L., Visscher, P. M., O’Donovan, M. C., Sullivan, P. F., … Consortium, T. I. S. (2009). Common polygenic variation contributes to risk of schizophrenia and bipolar disorder. Nature, 460(7256), 748–752. https://doi.org/10.1038/nature08185

Purves, K. L., Coleman, J. R. I., Meier, S. M., Rayner, C., Davis, K. A. S., Cheesman, R., … Eley, T. C. (2020). A major role for common genetic variation in anxiety disorders. Molecular Psychiatry, 25(12), 3292–3303. https://doi.org/10.1038/s41380-019-0559-1

Reed, Z. E., Micali, N., Bulik, C. M., Smith, G. D., & Wade, K. H. (2017). Assessing the causal role of adiposity on disordered eating in childhood, adolescence, and adulthood: A Mendelian randomization analysis. American Journal of Clinical Nutrition, 106(3), 764– 772. https://doi.org/10.3945/ajcn.117.154104

Ripke, S., Neale, B. M., Corvin, A., Walters, J. T. R., Farh, K.-H., Holmans, P. A., … Consortium, S. W. G. of the P. G. (2014). Biological insights from 108 schizophrenia-associated genetic loci. Nature, 511(7510), 421–427. https://doi.org/10.1038/nature13595

Robinson, L., Zhang, Z., Jia, T., Bobou, M., Roach, A., Campbell, I., … Desrivières, S. (2020). Association of Genetic and Phenotypic Assessments With Onset of Disordered Eating Behaviors and Comorbid Mental Health Problems Among Adolescents. JAMA Network Open, 3(12), e2026874. https://doi.org/10.1001/jamanetworkopen.2020.26874

Scott, R. A., Scott, L. J., Mägi, R., Marullo, L., Gaulton, K. J., Kaakinen, M., … Prokopenko, I. (2017). An Expanded Genome-Wide Association Study of Type 2 Diabetes in Europeans. Diabetes, 66(11), 2888–2902. https://doi.org/10.2337/db16-1253

Shisslak, C. M., Renger, R., Sharpe, T., Crago, M., McKnight, K. M., Gray, N., … Taylor, C. B. (1999). Development and evaluation of the McKnight risk factor survey for assessing potential risk and protective factors for disordered eating in preadolescent and adolescent girls. International Journal of Eating Disorders, 25(2), 195–214. https://doi.org/10.1002/(SICI)1098-108X(199903)25:2<195::AID-EAT9>3.0.CO;2-B

Solmi, F., Mascarell, M. C., Zammit, S., Kirkbride, J. B., & Lewis, G. (2019). Polygenic risk for schizophrenia, disordered eating behaviours and body mass index in adolescents. British Journal of Psychiatry, 215(1), 428–433. https://doi.org/10.1192/bjp.2019.39

Stahl, E. A., Breen, G., Forstner, A. J., McQuillin, A., Ripke, S., Trubetskoy, V., … Sklar, P. (2019). Genome-wide association study identifies 30 loci associated with bipolar disorder. Nature Genetics, 51(5), 793–803. https://doi.org/10.1038/s41588-019-0397-8

Stice, E. (1998). Modeling of eating pathology and social reinforcement of the thin-ideal predict onset of bulimic symptoms. Behaviour Research and Therapy, 36(10), 931–944. https://doi.org/10.1016/S0005-7967(98)00074-6

Stice, E. (2001). A prospective test of the dual-pathway model of bulimic pathology: Mediating effects of dieting and negative affect. Journal of Abnormal Psychology, 110(1), 124–135. https://doi.org/10.1037/0021-843X.110.1.124

Taylor, A. E., Jones, H. J., Sallis, H., Euesden, J., Stergiakouli, E., Davies, N. M., … Tilling, K. (2018). Exploring the association of genetic factors with participation in the Avon Longitudinal Study of Parents and Children. International Journal of Epidemiology, 47(4), 1207–1216. https://doi.org/10.1093/ije/dyy060

Teslovich, T. M., Musunuru, K., Smith, A. V., Edmondson, A. C., Stylianou, I. M., Koseki, M., … Kathiresan, S. (2010). Biological, clinical and population relevance of 95 loci for blood lipids. Nature, 466(7307), 707–713. https://doi.org/10.1038/nature09270

Tingley, D., Yamamoto, T., Hirose, K., Keele, L., & Imai, K. (2014). mediation : R Package for Causal Mediation Analysis. Journal of Statistical Software, 59(5), 1–38. https://doi.org/10.18637/jss.v059.i05

VanderWeele, T. (2015). Explanation in causal inference: methods for mediation and interaction. Oxford University Press.

Wade, T. D., Gordon, S., Medland, S., Bulik, C. M., Heath, A. C., Montgomery, G. W., & Martin, N. G. (2013). Genetic variants associated with disordered eating. International Journal of Eating Disorders, 46(6), 594–608. https://doi.org/10.1002/eat.22133

Walters, R. K., Polimanti, R., Johnson, E. C., McClintick, J. N., Adams, M. J., Adkins, A. E., … Agrawal, A. (2018). Transancestral GWAS of alcohol dependence reveals common genetic underpinnings with psychiatric disorders. Nature Neuroscience, 21(12), 1656– 1669. https://doi.org/10.1038/s41593-018-0275-1

Watson, H. J., Yilmaz, Z., Thornton, L. M., Hübel, C., Coleman, J. R. I. I., Gaspar, H. A., … Bulik, C. M. (2019). Genome-wide association study identifies eight risk loci and implicates metabo-psychiatric origins for anorexia nervosa. Nature Genetics, 51(8), 1207–1214. https://doi.org/10.1038/s41588-019-0439-2

Witt, S. H., Streit, F., Jungkunz, M., Frank, J., Awasthi, S., Reinbold, C. S., … Rietschel, M. (2017). Genome-wide association study of borderline personality disorder reveals genetic overlap with bipolar disorder, major depression and schizophrenia. Translational Psychiatry, 7(6), e1155–e1155. https://doi.org/10.1038/tp.2017.115

Wray, N. R., Ripke, S., Mattheisen, M., Trzaskowski, M., Byrne, E. M., Abdellaoui, A., … Sullivan, P. F. (2018). Genome-wide association analyses identify 44 risk variants and refine the genetic architecture of major depression. Nature Genetics, 50(5), 668–681. https://doi.org/10.1038/s41588-018-0090-3

Yengo, L., Sidorenko, J., Kemper, K. E., Zheng, Z., Wood, A. R., Weedon, M. N., … Visscher, P.M. (2018). Meta-analysis of genome-wide association studies for height and body mass index in ∼ 700000 individuals of European ancestry. Human Molecular Genetics, 27(20), 3641–3649. https://doi.org/10.1093/hmg/ddy271

