## Supplemental Figure S1 for "Eating disorder symptoms and their associations with anthropometric and psychiatric polygenic scores"

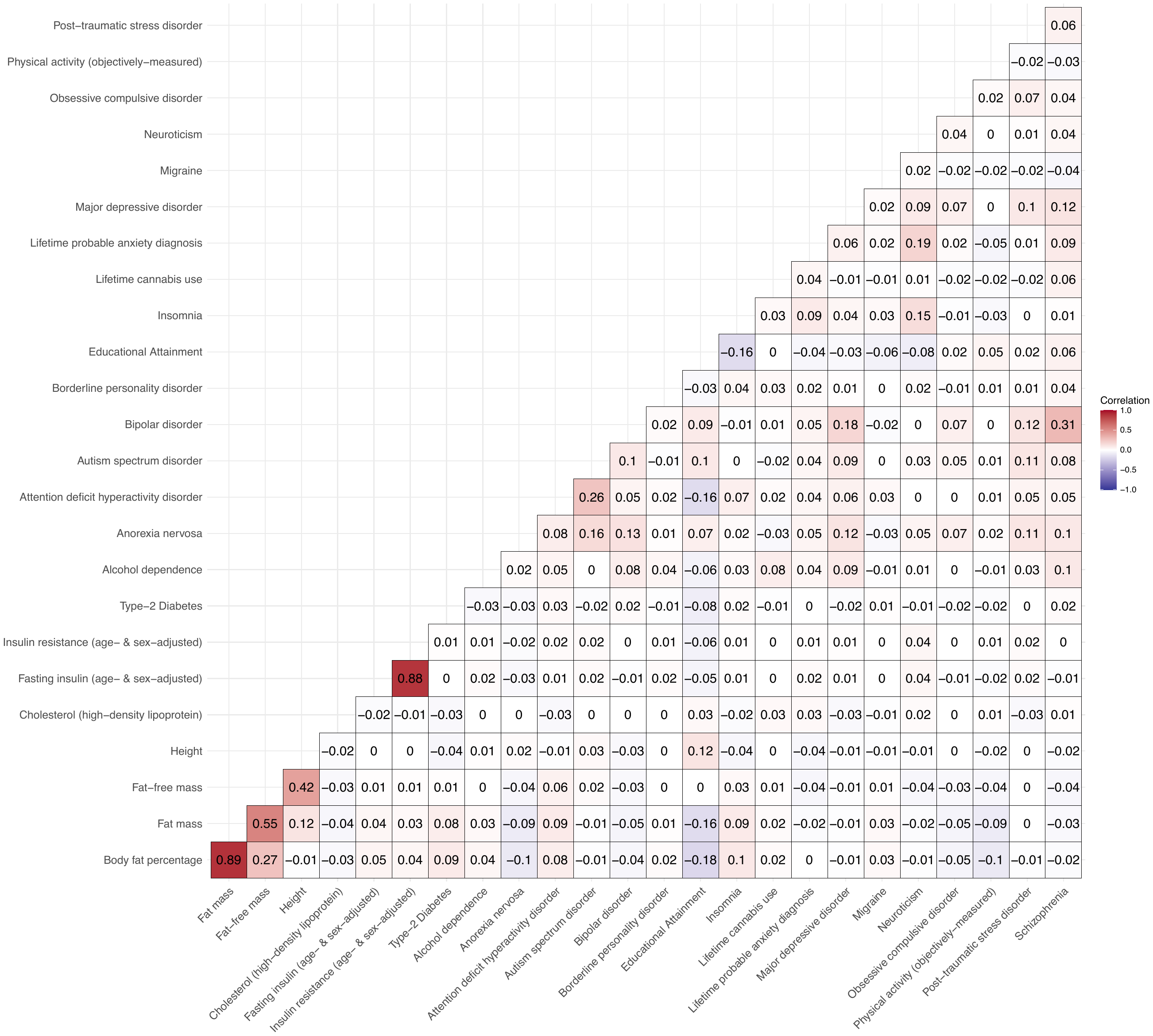

**Figure S1:** Correlation matrix of the investigated polygenic scores in the Avon Longitudinal Study of Parents and Children (ALSPAC) (Boyd et al., 2013; Fraser et al., 2013; Golding, 2004). For more information regarding sources of the polygenic scores see Table S1.

**References**

Boyd, A., Golding, J., Macleod, J., Lawlor, D. A., Fraser, A., Henderson, J., Molloy, L., Ness, A., Ring, S., & Smith, G. D. (2013). Cohort profile: The ’Children of the 90s’-The index offspring of the avon longitudinal study of parents and children. *International Journal of Epidemiology*, *42*(1), 111–127. https://doi.org/10.1093/ije/dys064

Fraser, A., Macdonald-wallis, C., Tilling, K., Boyd, A., Golding, J., Davey smith, G., Henderson, J., Macleod, J., Molloy, L., Ness, A., Ring, S., Nelson, S. M., & Lawlor, D. A. (2013). Cohort profile: The avon longitudinal study of parents and children: ALSPAC mothers cohort. *International Journal of Epidemiology*, *42*(1), 97–110. https://doi.org/10.1093/ije/dys066

Golding, J. (2004). The Avon Longitudinal Study of Parents and Children (ALSPAC)--study design and collaborative opportunities. *European Journal of Endocrinology*, U119–U123. https://doi.org/10.1530/eje.0.151u119
